# Evaluation of a serum protein signature as monitoring biomarker for Duchenne Muscular Dystrophy in a long-term clinical trial with corticosteroids

**DOI:** 10.64898/2025.12.18.25342544

**Authors:** Chiara Degan, Rebecca A. Tobin, Sharon I. de Vries, Albert Jiménez-Requena, Amela Peco, Michela Guglieri, Jordi Diaz-Manera, Yuri E. M. van der Burgt, Bart J. M. Vlijmen, DMD investigators of the Muscle Study Group, Yetrib Hathout, Cristina Al-Khalili Szigyarto, Utkarsh J. Dang, Roula Tsonaka, Pietro Spitali

## Abstract

**Objective:** Duchenne muscular dystrophy (DMD) is a progressive neuromuscular disorder for which monitoring biomarkers are urgently needed. We aimed to evaluate whether proteins in serum can accurately monitor patients’ function within the duration of a clinical trial.

**Methods:** In this study, we evaluated longitudinal serum proteins of DMD patients participating to the FOR-DMD clinical trial, comparing daily and intermittent corticosteroid regimens in boys aged 4-8 years at baseline. Using the aptamer-based protein platform SomaScan, we profiled 1500 proteins. Associations between protein levels and motor function outcomes, such as Rise from the Floor Velocity (RFV), 10-Meter Run/Walk Velocity (10MRWV), and North Star Ambulatory Assessment (NSAA), were assessed using linear mixed models. In particular, we explored whether patients with higher protein levels also tended to have better functional scores (across-patients analysis), and whether changes in protein levels within the same patient over time were linked to changes in their functional performance (within-patient analysis). Finally, penalized (Lasso) mixed models were applied to evaluate the predictive function of the proteins. The prediction accuracy of the models (evaluated by optimism-corrected Root Mean Squared Error) was compared to that of a simpler model with only age and treatment as predictors.

**Results:** Across-patients and within-patient analyses revealed consistent associations with three functional tests for a subset of proteins, notably RGMA, ART3, ANTXR2, and CFB. Multivariate models incorporating the proteins significantly associated with at least two tests, improved prediction accuracy for NSAA and RFV by 21% and 8%, respectively. These models also revealed a subset of proteins that were consistently selected. Quantification of CFB, RGMA, ANTXR2, SERPINF1 and ATP5PF using SomaScan showed strong agreement with measurements obtained using orthogonal methods such as ELISA, MRM-MS and an in-house developed bead-based sandwich immunoassay.

**Discussion:** These findings support the utility of serum protein signatures as objective, quantitative tools for monitoring disease progression and treatment response in DMD during clinical visits and clinical trials.

## Introduction

Duchenne Muscular Dystrophy (DMD) is a rare, progressive neuromuscular disorder caused by pathogenic variants in the *DMD* gene, resulting in loss of dystrophin expression. Patients typically exhibit delayed motor development, muscle weakness, progressive loss of muscle mass and mobility/ambulation, and reduced life expectancy. Motor decline is commonly assessed using clinical tools like the North Star Ambulatory Assessment (NSAA), and/or timed functional tests like Rise from the Floor Velocity (RFV), or 10-Meter Run/Walk Velocity (10MRWV).

While commonly used in interventional clinical trials, functional scores are inherently variable both within^1^ and across^2^ patients and often subjective, being influenced by patient motivation and collaboration, and evaluator’s ability to engage the patient. Moreover, these assessments provide limited insight into disease trajectories during the early years of life (ages 0–6), a critical period before significant muscle tissue replacement by adipose tissue occurs, and where these motor assessments may be less reliable^3^. These limitations challenge early-phase clinical trials, where therapeutic interventions may be most effective.

To address these challenges, researchers have explored alternatives for monitoring patient performance, including wearable technologies (e.g., 95th percentile stride velocity^4^), MRI-based fat quantification^5^, and biomarkers in body fluids^6^. Several studies identified protein biomarkers capable of distinguishing DMD patients from healthy controls using serum or plasma samples^7–10^, but only a few investigated longitudinal changes in these biomarkers, typically relying on retrospectively collected natural history cohorts^11^. However, retrospective studies are often confounded by factors including heterogeneous age ranges, variations in standards of care, differences in corticosteroid type, regimen and crossovers, and the inherent variability of natural history data compared to controlled clinical trials.

Monitoring biomarkers are objective, quantitative measures that can be used to assess disease progression and/or response to intervention. They may reflect changes that correlate with the worsening, stabilization, or improvement of a pathological condition over time. They may also serve as pharmacodynamic biomarkers by providing longitudinal information regarding the efficacy and safety of a given treatment. In the long run, biomarkers may even substitute clinical endpoints. However, developing accurate, sensitive, and reliable monitoring biomarkers is often challenged within research by the technological platforms employed, the biological variability between studied individuals, and the limited longitudinal data, especially in the context of rare diseases as DMD^12,13^. Finally, while single biomarkers have been mostly used to describe DMD vs. control differences^14^, response to treatments^15^ (e.g. corticosteroids), and associations with clinical outcomes^6^, panels of different biomarkers might improve the potential to monitor patients disease state and prediction of longitudinal outcomes.

In this study, we aim to bridge these gaps by analyzing longitudinal serum protein trajectories in relation to clinical performance using samples and clinical data from the FOR-DMD trial, a well-controlled cohort, consisting of intermittent prednisone, daily prednisone, and daily deflazacort groups^16^. We demonstrate that a panel of serum proteins associates with functional performance. We further show that in this cohort, this panel of proteins improves the prediction of performance metrics and functional decline over a 3-year follow-up period, compared to models without the biomarker data.

## Material and Methods

### FOR-DMD clinical trial and samples

The FOR-DMD study was registered at ClinicalTrials.gov (no. NCT01603407). Serum samples and clinical data were collected from 2013 to 2019, following approval from the competent ethics committee at each institution in accordance with the International Conference on Harmonisation guidelines for Good Clinical Practice and the World Medical Association Declaration of Helsinki. Informed written consent for participation in biomarker research was obtained from the parents or legal guardians of participants at the time of enrollment in the clinical study.

The study included biosamples and clinical outcomes on participants from the FOR-DMD clinical trial, treated with daily dosing of either deflazacort or prednisone, or intermittent dosing of prednisone^16^. Clinical assessments to monitor patients’ performance included Rise from the Floor Velocity (RFV), North Star Ambulatory Assessment (NSAA), and 10-Meter Run/Walk Velocity (10MRWV). RFV measures the speed at which a patient stands from a supine position.

Peripheral blood samples were collected using standard venipuncture techniques into serum separator tubes. Samples were clotted at room temperature for 30 minutes, then centrifuged at 1500g for 10 minutes at 4°C. The resulting serum was aliquoted into cryovials and stored at −80°C until analysis.

### Biomarkers selection

A custom panel of 1500 protein targets was defined for Somascan quantification. This list was compiled from evidence obtained from previously published biomarker datasets, biomarker curation with relevance to pathophysiology and progression, and exploratory clinical data analysis. Additional proteins were included for their known links to DMD genetic modifiers, exercise, metabolic syndrome, bone fragility, and genes expressed at high levels specifically in adipose tissue, heart, and brain^9–11,15,17–19^.

### SomaScan assay

Serum proteome profiling was performed using the SomaScan® Assay (SomaLogic, Inc.; Boulder, CO, USA), a high-throughput aptamer-based platform capable of exploring thousands of proteins simultaneously. The assay utilizes SOMAmer® (Slow Off-rate Modified Aptamer) reagents, which are chemically modified single-stranded DNA aptamers with high affinity and specificity for protein targets. For this study, a subset of the SomaScan 7K Assay version was used based on the selected biomarkers. Briefly, 55 µL of serum was incubated with a mixture of SOMAmer reagents. Protein-SOMAmer complexes were captured on streptavidin-coated beads, washed to remove unbound proteins, and then photocleaved to release SOMAmers. The released SOMAmers were quantified using hybridization to custom DNA microarrays, and signal intensities were measured via fluorescence scanning. Raw fluorescence intensity data were normalized using SomaLogic standard hybridization normalization, median signal normalization, and calibration procedures to correct for systematic biases and inter-plate variability. Quality control metrics (QC) assessed sample integrity, hybridization efficiency, and overall assay performance^20^; four samples failing QC thresholds were excluded from downstream analyses.

### Data pre-processing

Protein levels were log2-transformed and filtered based on several criteria: we removed proteins that had clear bimodality within DMD patients at baseline (possibly due to epitopes, SNPs, free or bound forms of proteins, complex formation, etc.), those that had too little variability, those proteins that were flagged by Somalogic quality check, and low-abundant proteins out of pairs of proteins with high correlations (> 0.90). We assessed the correlation by accounting for the longitudinal nature of the proteins. We first modeled each log2-protein over time using linear mixed-effects models (LMMs) with random intercepts and age included as a fixed effect. We then calculated Spearman correlations between the model-predicted protein levels. The correlation was estimated at the median age of 7.18, where the data were most densely concentrated.

To assess the presence of batch effects in Somascan profiles, we used a principal component analysis (PCA) on the log2-transformed proteins.

### Statistical analysis

Associations between serum protein levels and functional performance were assessed using LMMs. Separate LMMs were fitted for each functional outcome. Each included fixed effects for age, treatment group, log2-transformed protein level, the interaction between age and treatment group (allowing treatment effects to vary with age), and between protein level and treatment group (allowing protein levels to vary with treatment effects). Random intercepts were included to account for within-subject correlation due to repeated measures. This approach allowed evaluation of the relationship between protein levels and functional performance both across patients and within patients over time. For the latter, LMMs have been applied to the within-patient changes, which were computed by centering each patient’s observations (both function scores and proteins) around their mean across time. Associations were tested using the F-test, and the p-values across all proteins have been corrected for multiple testing using the FDR method. Proteins significantly associated (FDR-adjusted p-value < 0.05) with at least two longitudinal functional tests were retained for further analysis.

To assess predictive capability of multiple proteins simultaneously, we used penalized mixed-effects models with an L1-penalty (Lasso) on protein fixed effects (log2-transformed and standardized). This approach enable variable selection in the presence of repeated measurements per subject. Treatment and age were also included in the model, but without penalization. The L1-penalty depends on a tuning parameter, which determines how many proteins the model selects. To optimize this parameter, we used subject-specific cross-validation (5-fold), ensuring no data from the subjects on the test set was used for training. Model performance was evaluated using root mean squared error (RMSE). RMSE quantifies the average discrepancy between the observed and predicted values of the functional score, and provides a measure of model fit in the original outcome scale. We employed internal validation using a bootstrap procedure^21^. For each bootstrap sample (out of the 1000 bootstrap samples used), we fit a cross-validated lasso model and computed the RMSE in both the bootstrap sample and the original dataset. The RMSE difference quantified overfitting (optimism), which we averaged across all samples. Subtracting this average optimism from the original RMSE gave an optimism-corrected RMSE^21^. Finally, we evaluated the model performance by comparing the RMSE of the protein-based model to that of a null model including only age and treatment group as predictors. The relative difference in RMSE quantifies the improvement/degradation in predictive accuracy attributable to the inclusion of the selected proteins. We also report the frequency of each protein selection across the 1000 repeated analyses. This metric, referred to as Variable Inclusion Probability^22^ (VIP), provides an estimate of the likelihood that a protein is included in the model rather than excluded, and offers insight into model stability and the robustness of feature selection. LMMs were implemented using the R packages *lme4*^23^ and *lmerTest*^24^, and penalized mixed models using the R package *glmmLasso*^25^.

### Sample set for orthogonal validation

Orthogonal assays were used to validate Somascan signals, employing methods such as multiple reaction monitoring by mass spectrometry (MRM-MS), ELISA, and suspension bead immunoassay.

Orthogonal validation by sandwich immunoassay required only 6 μl of serum and was performed on the same FOR-DMD samples used for protein trajectory analysis.

Due to limited sample volume in the FOR-DMD cohort, MRM-MS and ELISA validations were performed using 80 independent serum samples from a retrospective study involving DMD patients followed at Leiden University Medical Center (LUMC) between 2009 and 2022^26^, for which Somascan data were available. These serum samples were obtained from 17 individuals aged 4.8 to 18.7 years, with 2 to 7 observations per patient (mean: 4.7), during routine annual outpatient clinic visits. Serum was prepared following standard phlebotomy procedures: samples were clotted at room temperature for approximately 30 minutes and subsequently centrifuged at 2350g for 10 minutes. Written informed consent was obtained from all participants or their legal guardians in accordance with protocol B22.013, approved by the regulatory board at LUMC, called "*Medische Ethische Toetsings Commissie*" (METC, medical ethics review committee).

### Validation with orthogonal methods

Targeted quantification of SERPINF1 and CFB was performed using an in-house developed MRM assay that included internal standard peptides for each protein of interest^27^. Serum proteins were denatured, alkylated, reduced, and digested into quantifier tryptic peptides following established protocols^28,29^. The MRM assay was developed according to Tier 2 guidelines^30,31^ and optimized for serum analysis. Protein quantities were reported as ratios between endogenous quantifier peptides and heavy-labelled internal standard peptides spiked into the samples at known amounts. The analytical performance was monitored using pooled plasma QC samples from healthy donors.

Candidate biomarkers like RGMA, ART3, and ANTXR2 that are not yet included in the MRM assay were assessed using ELISA. RGMA and ANTXR2 were analyzed in 76 serum samples using DuoSet ELISA kits (Bio-Techne #DY2459-05 and DY2940-05, respectively) according to manufacturer’s protocol. Briefly, 10-13 µl (RGMA) or 10 µl (ANTXR2) of serum was processed in duplicate and measured using an ID3 plate reader at 450 nm, with a correcting wavelength at 570 nm. For RGMA, 28 samples were excluded from the analysis due to undetectable levels (n=20) or high Coefficient of Variation (CV) (n=8). Concentrations in pg/ml were calculated after averaging and interpolating the log-transformed values with a Four Parameter Logistic (4PL) regression. CVs did not exceed 11.9% (RGMA) or 13.5% (ANTXR2), with mean CVs of 4.3% and 2.1%, respectively. ART3 was assessed with ELISA (Novus Biologicals, #NBP2-69866) in 5 samples according to manufacturer’s protocol. Serum samples were tested using different dilutions ranging from 1:4 to 1:64.

For ATP5PF, a self-sandwich immunoassay was developed using a suspension bead array and a validated anti-ATP5PF monospecific antibody^32^ (HPA031069). The anti-ATP5PF antibody was coupled to MagPlex microspheres, to be used as capture antibody according to previously established protocols^33^ with minor changes. Two negative controls were included to assess the background, one with beads not coupled to antibodies and a second one coupled with 17.5 µg/ml rabbit serum immunoglobulin G. Subsequent coupling with antibodies, the beads were blocked with 50 µl of 5% (w/v) Blocking Reagent for nucleic acid hybridization and detection (Roche) prepared in PBST. Six µg of anti-ATP5PF antibodies were separately biotinylated to be used as detection antibody according to previously established protocol^33^. Quantification of ATP5PF was performed using the sandwich immunoassay constructed and the protein epitope signature tag (PrEST) antigen (APrEST86637) from Atlas Antibodies AB as protein standard^34^. The ATP5PF PrEST was diluted to 17 different concentrations between 0.039 and 140 ng/ml in assay buffer supplemented with the corresponding amount of guinea-pig serum to create the standard curve. Serum samples, more specifically, 6 µl were diluted in assay buffer containing 0.025% (w/v) polyvinyl alcohol, 0.04% (w/v) polyvinylpyrrolidone (Sigma), 0.005% (w/v) casein (Sigma), 0.5 mg/ml rabbit IgG, and 1:1000 ProClin™ 300 in PBST, up to 50 µl and analysed^33^. The assay was read on a Luminex® FlexMap 3D™ system (Luminex Corp.) using xPONENT software, and median fluorescence intensities (MFI) were recorded for all samples and standards. Standard curves were fitted using a five-parameter log-logistic regression model^35^. For each plate, the linear working range was defined as the interval between two standard deviations above the mean background MFI and 90% of the MFI of the highest standard. The MFI were normalized using probabilistic quotient normalization^36^ to remove the batch effect between plates.

### Data Availability

Anonymized data can be made available to qualified investigators on request. Requests should be in line with the approved ethical protocol.

## Results

### Clinical trajectories in the FOR-DMD cohort

The FOR-DMD study reported that daily treatment with either deflazacort or prednisone was more effective than intermittent treatment^16^. The original dataset included 196 participants aged 4–8 years at baseline (mean: 5.85, SD: 0.99), randomized to receive daily prednisone (n=65, average baseline age of the group: 5.86) dosed at 0.75 mg/kg/d, daily deflazacort (n=65, average baseline age of the group: 5.75) dosed at 0.90 mg/kg/d, or intermittent 10 days on/10 days off prednisone (n=66, average baseline age of the group: 5.94) dosed at 0.75 mg/kg/d. Across the full cohort, 1444 visits were recorded at baseline 3, 6, 12, 18, 24, 30, and 36 months.

For the current analysis, a subset of 56 patients was selected matching the availability of serum samples. Serum samples were only collected at baseline, 12, 24, and 36 months, resulting in 149 clinical visits that could be matched to sampling timepoints. Among the selected patients, one serum sample was obtained from 12 patients, whereas 44 patients had two or more. The baseline average age was 5.86 years (min: 4.24, max: 7.83). Of these 56 patients, 19 received daily prednisone (average baseline age of the group: 5.98), 14 daily deflazacort (average baseline age of the group: 5.45), and 23 intermittent prednisone (average baseline age of the group: 6.00).

For the analysis, we considered three outcomes: Rise from the Floor Velocity (RFV), 10-Meter Run/Walk Velocity (10MRWV), and North Star Ambulatory Assessment (NSAA). Table 1 summarizes and compares baseline scores between the full FOR-DMD dataset and the selected subset. Modeling of the original data confirmed that patients treated daily were performing better in all three tests compared to patients treated intermittently (RFV and 10MRW: p-values < 0.001; NSAA: p-value = 0.003). Instead, analysis of the non-randomized subset showed that daily deflazacort treated patients were performing better on the RFV and 10MRWV compared to daily prednisone and intermittent prednisone treated groups (RFV: p-value = 0.05; 10MRW: p-value = 0.02). This difference was partly explained by the younger baseline age in the daily deflazacort group (Figure 1A).

**Figure 1.**
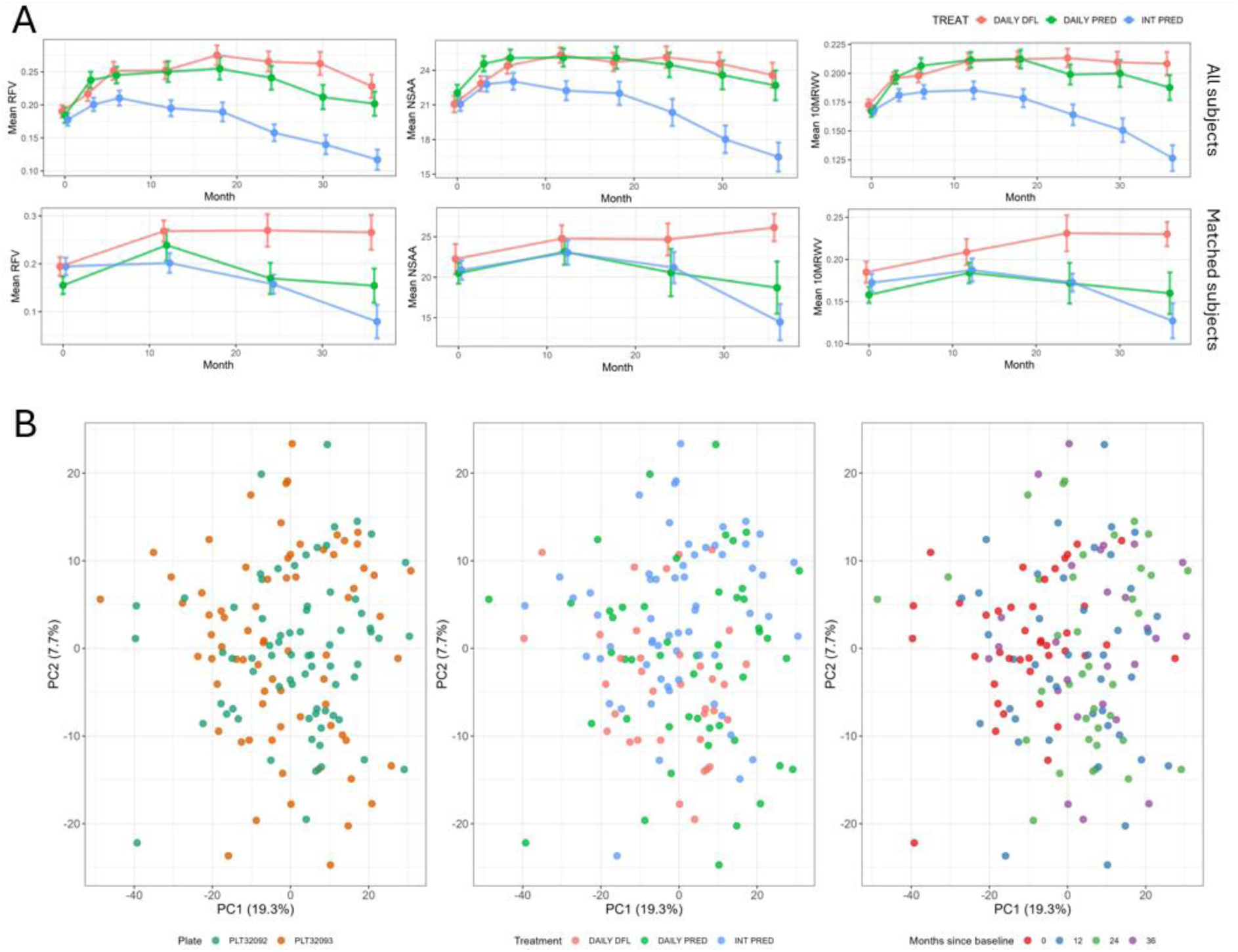
(A) Means and standard errors of monthly patient trajectories for NSAA, RFV, and 10MRWV shown for both the full FOR-DMD dataset and the subset with matched serum samples. (B) First and second principal components of the PCA (and explained variance) on proteins measured with Somalogic and selected after preprocessing, stratified by plate, treatment group, and visit time. NSAA: North Star Ambulatory Assessment; RFV: rise from the floor velocity; 10MRWV: 10-m run/walk velocity; PCA: Principal Component Analysis; PC1 and PC2: first and second Principal Components; DFL: deflazacort; PRED: prednisone.

**Table 1.** Descriptive statistics of baseline characteristics (mean, standard deviation, and median) for RFV, 10MRWV, and NSAA functional tests, shown for the full dataset and for the subset of measurements matched to serum samples.

| Treatment group | RFV* | 10MRWV* | NSAA* |
| --- | --- | --- | --- |
| *All participants* |  |  |  |
| DAILY PRED<br>(n=65) | 0.1839 (0.09272);<br>0.1667 | 0.1668 (0.03892);<br>0.1667 | 21.90 (5.545);<br>21 |
| DAILY DFL<br>(n=65) | 0.1896 (0.07433);<br>0.1754 | 0.1722 (0.03753);<br>0.1695 | 21.06 (5.780);<br>21 |
| INT PRED<br>(n=66) | 0.1793 (0.07367);<br>0.1754 | 0.1679 (0.03185);<br>0.1695 | 21.10 (4.956);<br>22 |
| *Matched to participants with at least 1 serum sample available* |  |  |  |
| DAILY PRED<br>(n=19) | 0.1652 (0.06518);<br>0.1667 | 0.1587 (0.03597);<br>0.1408 | 20.59 (5.066);<br>21 |
| DAILY DFL<br>(n=14) | 0.1781 (0.04928);<br>0.1754 | 0.1796 (0.03820);<br>0.1695 | 21.54 (5.485);<br>19 |
| INT PRED<br>(n=23) | 0.1858 (0.06421);<br>0.2041 | 0.1719 (0.03465);<br>0.1695 | 20.75 (4.362);<br>20 |
| NSAA: North Star Ambulatory Assessment, RFV: Rise from the Floor Velocity, 10MRWV: 10-Meter Run/Walk Velocity. DAILY PRED: daily prednisone, DAILY DFL: daily deflazacort, INT PRED: intermittent prednisone.<br>*Mean (SD); Median |  |  |  |

Pre-processing reduced the Somascan biomarker list from 1500 to 1251 proteins. PCA did not identify clear outliers nor a batch effect related to the running plate. There was no clear clustering of samples in relation to treatment arms and visits (Figure 1B).

### Association of serum proteins with clinical outcomes

Paired clinical and proteomic data enabled analyses of associations between protein levels and functional performance both across patients and within patients over time. Across-patient analysis aimed to identify associations between the levels of circulating proteins and the levels of functional outcomes. In contrast, within-patient analysis examined whether changes in protein levels per individual were associated with corresponding changes in their functional performance.

Analysis across patients identified 27 proteins associated with RFV, 5 with NSAA, and 24 with 10MRWV (Figure 2B). Four proteins were associated with all three outcomes, namely RGMA, ART3, ANTXR2, and CFB, while one protein (RARRES2) was associated with NSAA and 10MRWV, and seven proteins were associated with rise and run velocity (OXSM, ACOT12, ATP5PF, SERPINF1, SPA17, COQ7, PEX14).

**Figure 2.**
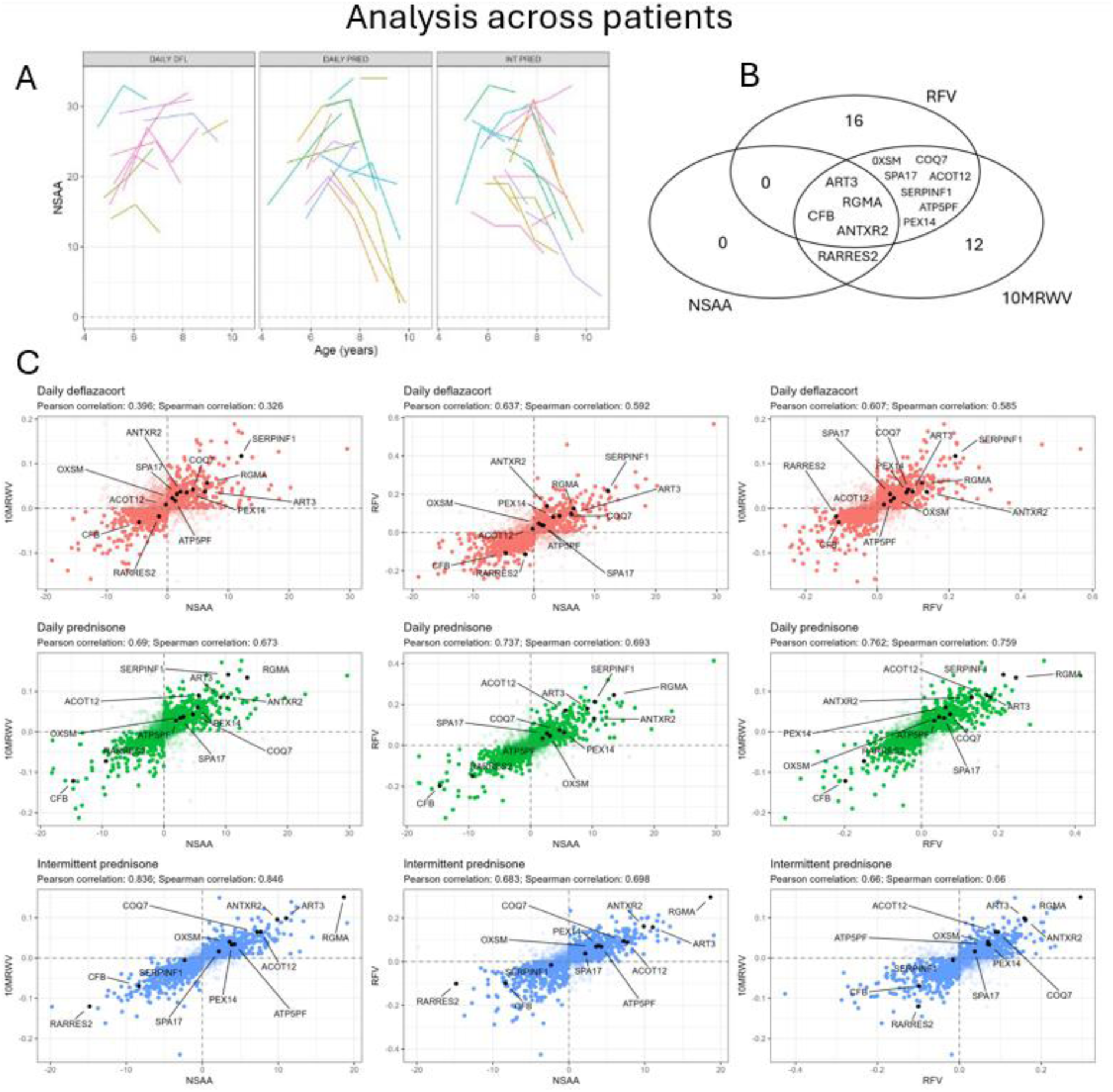
Results of the univariate analysis assessing associations between proteins and physical performance tests across patients. (A) Longitudinal trajectories of NSAA across age for all patients, grouped by treatment. Each line, distinguished by color, represents an individual patient. The data shown are in the original scale. (B) Venn diagram illustrating the name and number of proteins significantly associated with NSAA, 10MRWV, and RFV. (C) Scatterplots of protein effect estimates, stratified by physical test and treatment group. Colors indicate treatment groups: red for daily deflazacort, green for daily prednisone, and blue for intermittent prednisone. The 12 proteins associated with at least two physical tests are highlighted in black. NSAA: North Star Ambulatory Assessment; RFV: Rise from the Floor Velocity, 10MRWV: 10-Meter Run/Walk Velocity.

Associations were generally consistent across functional scores (Pearson R between 0.64 and 0.85, Figure 2C), except for NSAA vs. 10MRWV in the daily deflazacort group. Ten of the 12 proteins significantly associated with at least two outcomes showed consistent effects, both in direction and proportional magnitude, across tests and treatment arms. The two exceptions were ACOT12 and SERPINF1. ACOT12 levels were significantly and positively associated with 10MRWV and RFV in both prednisone treatment arms, but no clear association in the daily deflazacort group. Similarly, SERPINF1 showed a high positive association with 10MRWV and RFV, with the exception of patients in the intermittent prednisone group. Parameter estimates are provided in Table S1.

We then sought to determine whether changes in performance tests would be reflected in changes in protein levels. A group of 14 proteins (LEP, ART3, IL6, COQ7, OXSM, PFKM, RPIA, PEX14, NAMPT, CFB, ACOT12, PDHX, VSIG4, ANTXR2) significantly associated with within-patient changes in all three functional outcomes. In total, 61 proteins were associated with at least two functional tests. Notably, 11 of the 12 proteins associated with function across patients were found to be significant in the analysis within patients as well (all except RARRES2). A total of 36 proteins were associated with both 10MRWV and RFV (including RGMA), 4 with NSAA and RFV, and 6 with NSAA and 10MRWV. All three functional scores demonstrated comparable association patterns, showing correlation between 0.53 and 0.87 (Figure 3C). Proteins associated with functional scores both across and within patients showed consistent directional effects across treatment groups. As in the across-patient analysis, SERPINF1 remained significantly associated with RFV and 10MRWV, and showed a high positive association except under intermittent prednisone. Parameter estimates are provided in Table S2.

**Figure 3.**
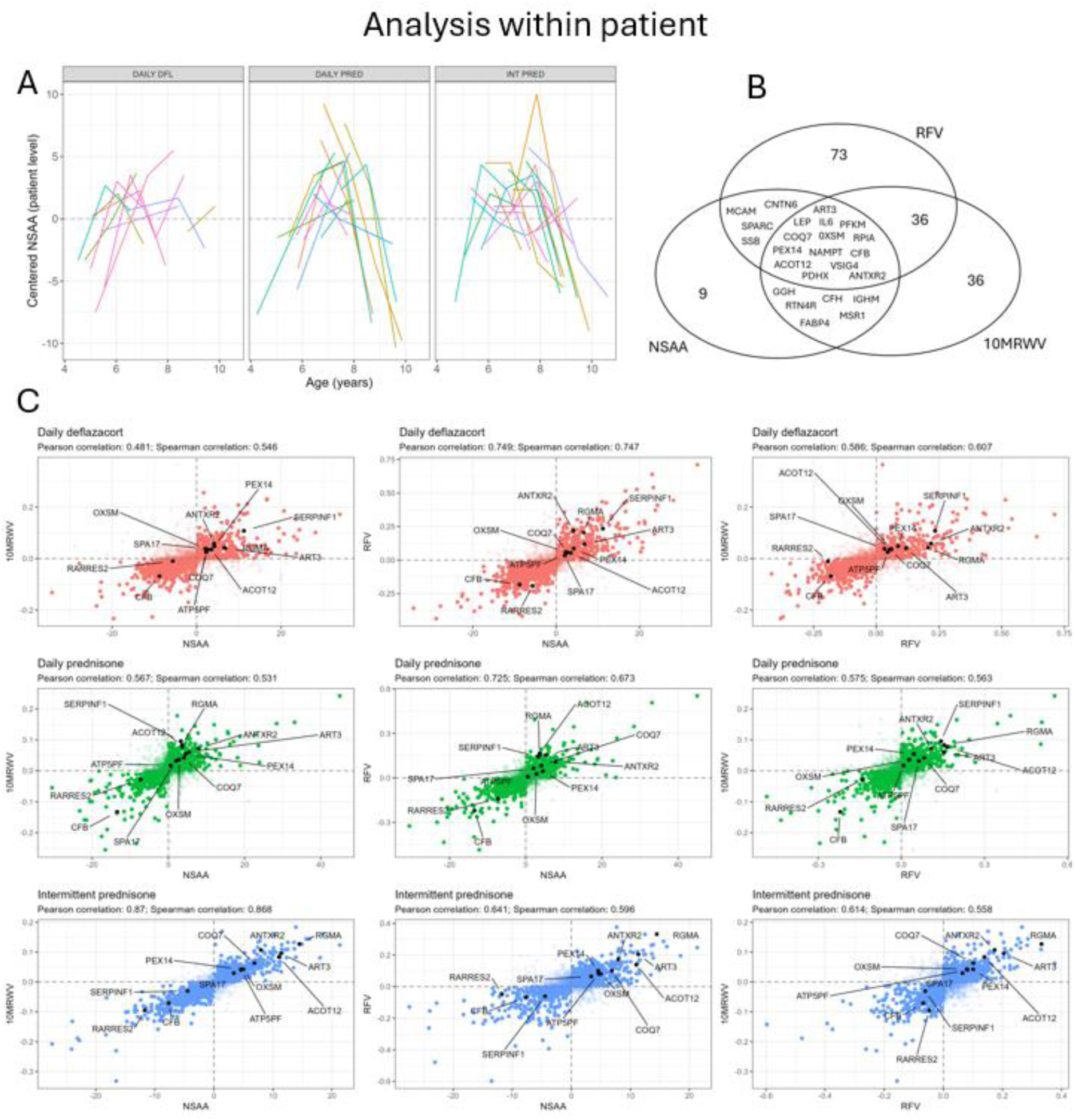
Results of the univariate analysis assessing associations between changes in proteins and changes in physical performance within-patients. (A) Longitudinal trajectories of NSAA across age for all patients, grouped by treatment. Each colored line represents an individual patient. Data are rescaled by subtracting each patient’s average value across all time points. (B) Venn diagram illustrating the name and number of proteins significantly associated with NSAA, 10MRWV, and RFV. (C) Scatterplots of protein effect estimates, stratified by physical test and treatment group. Colors indicate treatment groups: red for daily deflazacort, green for daily prednisone, and blue for intermittent prednisone. The 12 proteins associated with at least two physical tests both in across and within-patient analysis are highlighted. NSAA: North Star Ambulatory Assessment; RFV: Rise from the Floor Velocity, 10MRWV: 10-Meter Run/Walk Velocity.

Comparison of adjusted p-values in the analyses across and within patients supported the selection of the 4 proteins associated with all 3 functional scales for validation, namely RGMA, ART3, ANTXR2, and CFB (Figure S1).

### Predictive capability of the selected proteins

Given that 12 proteins were significant predictors of functional performance, and 61 proteins were significant predictors of change in function, we assessed whether combining these proteins into a panel could improve prediction accuracy of functional scores and score changes.

We initially applied Lasso penalization to the available dataset and assessed predictive performance by correlating observed and predicted outcome values. Strong correlations were observed for all three functional tests, both within and across patients (Figure 4).

**Figure 4.**
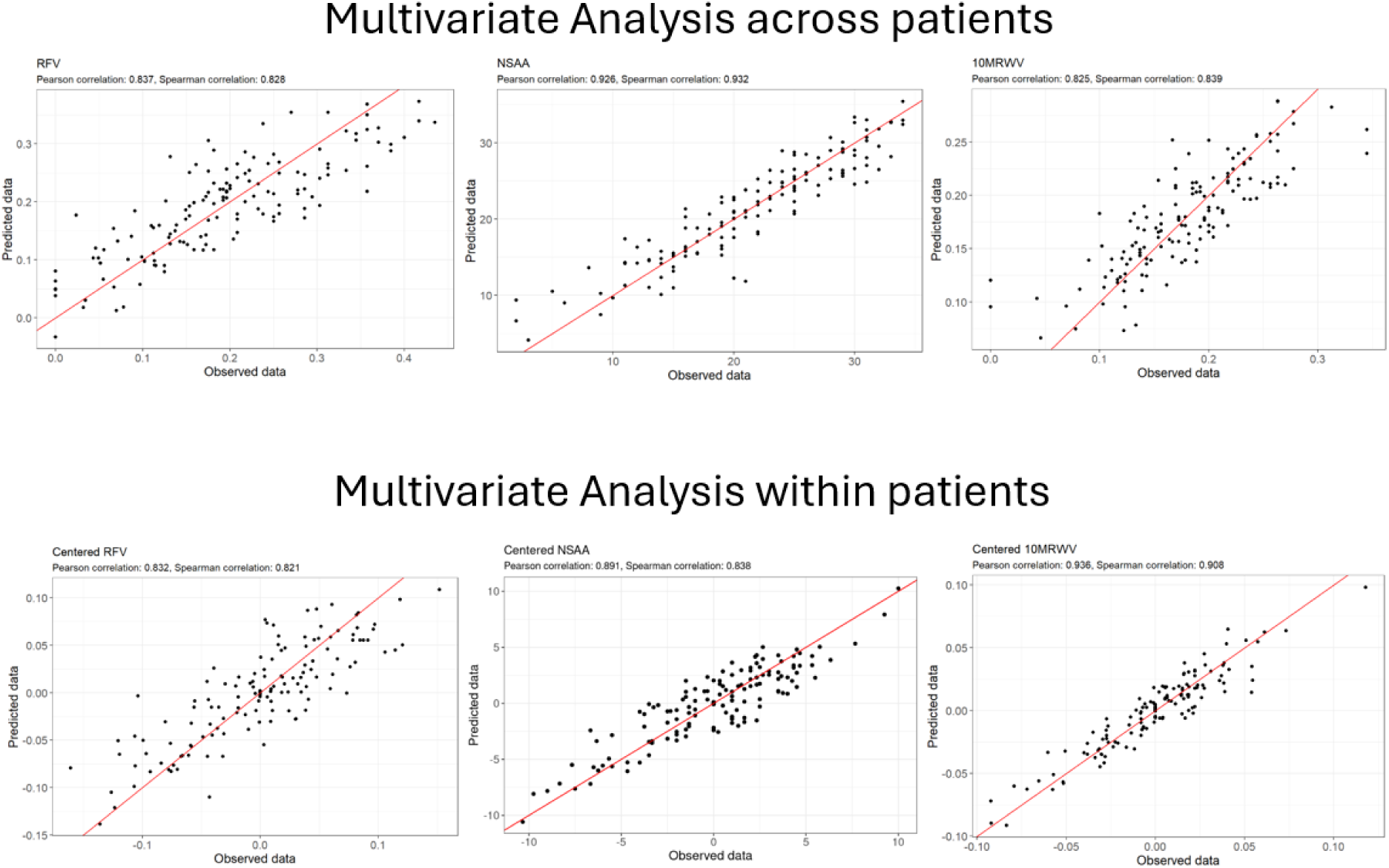
Scatterplots showing the correlation between the observed and predicted functional tests using multivariate models. ‘Observed’ refers to the actual measured values from the tests, while ‘predicted’ represents the values estimated by the model. Top panel: Comparison of predicted and observed data across-patients with a multivariate model including 12 proteins. Bottom panel: Comparison of predicted and observed data within-patients with a multivariate model including 61 proteins. NSAA: North Star Ambulatory Assessment; RFV: Rise from the Floor Velocity, 10MRWV: 10-Meter Run/Walk Velocity.

To validate these findings, we re-performed the analysis on 1000 random samples of the original dataset and assessed prediction accuracy using RMSE. Inclusion of the 12 proteins in the across-patient analysis improved predictive performance for NSAA and RFV by 21% and 8%, respectively (Table 2), while no improvement was observed for models aimed to predict the 10MRWV. For NSAA, the model achieved an RMSE of 3.03, indicating that predictions deviated from actual values by an average of approximately 3 points; an improvement over the null model, which had an RMSE of almost 4. The same analysis within patients further indicated that including proteins in the model reduced the prediction error by approximately 25–30% across all outcomes (Table 2).

**Table 2.** Evaluation of the model’s predictive performance in estimating functional test outcomes based on the 12 proteins across patients, and on the 61 proteins within patients. The RMSE of the model incorporating proteins is compared to that of a baseline model excluding proteins.

| Functional tests | RMSE null-model | RMSE protein-based model | Absolute difference | Relative difference* (%) |
| --- | --- | --- | --- | --- |
| *Across-patient analysis* |  |  |  |  |
| NSAA | 3.8576 | 3.0341 | 0.8235 | 21.36 |
| RFV | 0.0647 | 0.0592 | 0.0055 | 8.50 |
| 10MRWV | 0.0341 | 0.0352 | -0.0011 | -3.23 |
| *Within-patient analysis* |  |  |  |  |
| Within-patients NSAA changes | 3.6471 | 2.7245 | 0.9226 | 25.29 |
| Within-patients RFV changes | 0.0587 | 0.0398 | 0.0173 | 30.26 |
| Within-patients 10MRWV changes | 0.0316 | 0.0239 | 0.0077 | 24.37 |
| RMSE: Root Mean Squared Error, NSAA: North Star Ambulatory Assessment, RFV: Rise from the Floor Velocity, 10MRWV: 10-Meter Run/Walk Velocity. |  |  |  |  |
| *(RMSE null model - RMSE lasso)/(RMSE null model) * 100 |  |  |  |  |

We also used the 1000 repetitions to estimate the proportion of times each protein was selected and to assess the stability of these selections. In the across-patient analysis, 7 proteins (RGMA, ART3, COQ7, PEX14, ANTXR2, ATP5PF, and SPA17) out of the 12 proteins were retained in >90% of models (Table S3). When predicting change in function, two proteins (FCRL4 and SSB) were consistently excluded across all outcomes, while the remaining 59 were retained but at lower frequencies than in the across-patient analysis. Notably, the seven most frequently selected proteins in the across-patient analysis were also among the most stable in the within-patient models, with ART3, PEX14, COQ7, SPA17, and ANTXR2 showing the highest retention rates. All frequencies are summarized in Table S3.

### Orthogonal validation of candidate monitoring biomarkers

Because associations depend on the assays used to measure protein levels, we selected a list of candidate biomarkers for orthogonal validation using alternative assays. Proteins were selected based on their estimated effect across multiple functional assessments, their differential associations between treatment groups, and their retention frequency in multivariate models. Based on these criteria, CFB, SERPINF1, ART3, RGMA, ANTXR2 and ATP5PF were prioritized. CFB, ART3, and ANTXR2 were associated with all three functional scores across and within patients. ATP5PF was significantly associated with 10MRWV and RFV, as well as their longitudinal changes, and reached 100% VIP in the multivariate models. RGMA was the only protein retained in 100% of across-patient models. SERPINF1 was associated with functional performance across and within patients, with the exception of the intermittent prednisone group.

While the ART3 assay was in our hands not sufficiently sensitive to detect reduced ART3 levels in DMD patients, assays for RGMA, ANTXR2, CFB, SERPINF1 and ATP5PF showed strong concordance with Somascan measurements, supporting the validity of the findings (Figure 5). Somascan measurements show strong positive correlations with ELISA for RGMA and ANTXR2, with MRM-MS for SERPINF1 and CFB, and with suspension bead array immunoassay for ATP5PF, with Pearson coefficients ranging from 0.70 to 0.884 and Spearman coefficients from 0.728 to 0.902.

**Figure 5.**
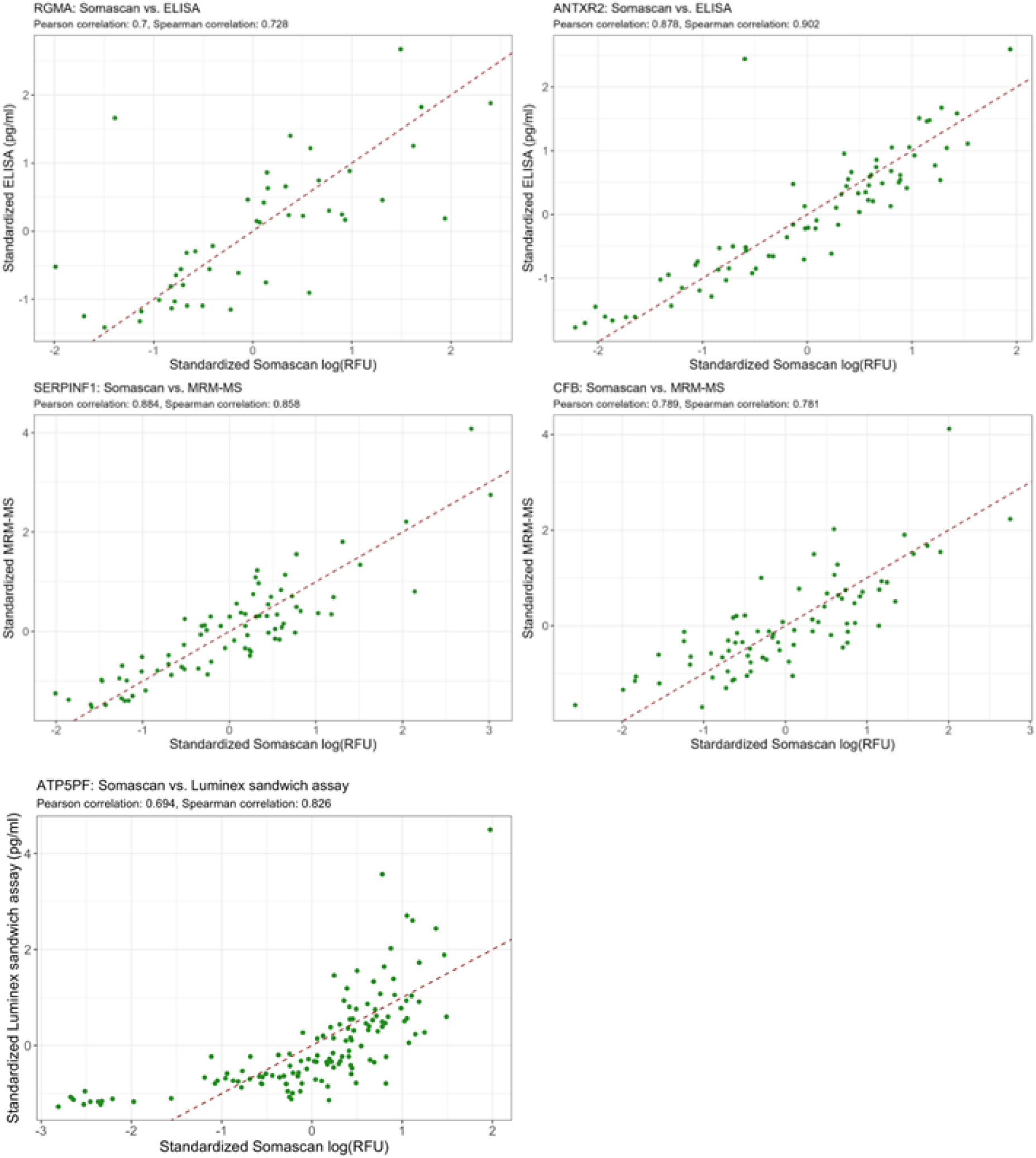
Orthogonal validation of selected proteins. Comparison between Somascan, ELISA, MRM-MS and an in-house developed bead-based sandwich immunoassay for RGMA, ANTXR2, SERPINF1, CFB and ATP5PF. Standardized values: (values – mean)/sd.

## Discussion

Accessible blood biomarkers are critically needed to monitor disease progression and treatment response in DMD, particularly in younger boys where functional outcomes remain subjective, and in older patients where such measures are unavailable. In this study, we analyzed 1500 SomaScan serum proteomic profiles alongside clinical data from a well-characterized cohort of DMD patients enrolled in the FOR-DMD trial^16^, identifying a set of candidate biomarkers for patients monitoring.

Our analysis revealed several proteins associated with key functional outcomes (RFV, 10MRWV, NSAA), and longitudinal changes in these proteins mirrored changes in functional scores. Multivariate modeling showed that a panel of proteins improved prediction accuracy for most outcomes in both across and within-patient analysis, with the only exception of 10MRMV, where the model without any protein performed slightly better than the one with proteins. Interestingly, more proteins were selected by the within-patient analysis. This suggests that a larger panel of proteins might be needed to be able to monitor progression, while a significantly smaller set of proteins is able to separate stronger and weaker patients.

Among all proteins identified, four stood out: ART3, ANTXR2, RGMA, ATP5PF and CFB.

ART3 (ecto-ADP-ribosyltransferase 3) has been reported to be decreased at the mRNA level in the skeletal muscle of boys with DMD^37,38^ and, more recently, at the protein level in serum of glucocorticoid-naïve DMD patients compared with age-matched healthy controls^10^. It was also found to be associated with functional scores in two large independent retrospective longitudinal cohorts of DMD patients^6^. Circulating ART3 is reduced in other neuromuscular diseases, including amyotrophic lateral sclerosis (ALS) and limb-girdle muscular dystrophy^39,40^. Although ART3 is expressed in both slow- and fast-twitch muscle fibers^41^, its precise role in muscle pathogenesis remains unclear. In this study, we show that ART3 positively correlates with all three clinical outcomes, meaning that DMD subjects with higher levels of circulating ART3 perform better than those with lower ART3 circulating levels. Changes in ART3 levels positively correlated with the changes in the functional tests, indicating that shifts in protein levels in a patient correspond to changes in the performance. Furthermore, ART3 was selected >90% of the times by the multivariate models predicting functional performances, and in 70-90% of the models predicting changes over time, underline its robustness as a candidate biomarker.

ANTXR2 showed a similar pattern, correlating with clinical outcomes both in this study and in previous retrospective cohorts^6^, and being reduced in serum of glucocorticoid-naïve DMD patients^10^. ANTXR2 expression is elevated in muscle tissue of DMD patients compared to healthy controls, and mainly expressed by Fibro-adipogenic Progenitors^41^. Its absence leads to collagen accumulation in skeletal muscle^42–44^. Like ART3, ANTXR2 was also found at lower levels in ALS^40^. In our study, ANTXR2 emerged as one of the most stable predictors, retained in over 90% of multivariate models for functional performance and in >65% of models predicting changes in RFV and 10MRWV.

RGMA likewise correlated with all clinical outcomes, but its circulating levels were not significantly different between glucocorticoid-naïve DMD patients and age-matched healthy controls^10^. RGMA was reported to induce myoblast cell fusion via interaction with myogenin^45^. In our study, RGMA stood out as the most consistently selected predictor, retained in 100% of across-patients multivariate models.

ATP5PF was found to be positively associated with motor performance, particularly with RFV and 10MRWV in both analysis across- and within-patients. ATP5PF is a subunit of the mitochondrial ATP synthase complex, ubiquitously expressed in all tissues, playing an important role in oxidative phosphorylation and cellular energy production. While ATP5PF itself has not been previously directly linked to DMD, mitochondrial dysfunction and impaired oxidative phosphorylation have been described as characteristics of the dystrophic muscle^46^, postulating ATP5PF as a biologically plausible candidate marker of muscle energetic status in the DMD context. In addition, elevated serum levels of ATP5PF and related ATP synthase complex subunits have been associated with coronary heart disease^47^.

CFB, on the other hand, was negatively correlated with all three clinical outcomes, meaning that DMD boys with high levels of circulating CFB performed worse than those with low CFB circulating levels. CFB is an activator of the alternative complement pathway, leading to membrane attack complex formation. Although its circulating levels were not significantly different between glucocorticoid-naïve DMD patients and age-matched healthy controls^10^, this association might suggest a role of alternative complement pathway in DMD muscle pathogenesis. This is consistent with previous findings where genetic ablation of complement C3 improved muscle health in other models of muscular dystrophy as dysferlin-deficient mice^48^. However, further studies are needed to elucidate the role of CFB in DMD muscle pathogenesis.

Our study identified associations between proteins and clinical outcomes that were not preserved across treatment groups. An example is SERPINF1, which was positively associated with function in patients treated daily with prednisone or deflazacort, but not in those on intermittent prednisone. This relationship was missed in previous biomarker studies due to the retrospective nature of the data, which did not enable to control for the corticosteroid type and regimen. Given the association with function and treatment, monitoring SERPINF1 could be used to adapt steroid dosing in patients to balance the positive and side effects.

Our results support the use of a panel of proteins to predict monitoring of patients. Such panel could enable more frequent assessments in further prospective studies via remote phlebotomy than in-person clinical visits may allow. However, using a protein panel complicates the development of quantitative assays for biomarker monitoring. In this study, we prioritized the set of proteins already mentioned as relevant (CFB, ART3, RGMA, ANTXR2, ATP5PF) and those showing differential response across treatment arms (SERPINF1) for validation purposes. Assays for CFB, RGMA, ANTXR2, ATP5PF and SERPINF1 confirmed that the relative quantification by SomaScan correlated with refined quantification methods as MRM-MS, ELISAs and suspension bead immunoassay, suggesting that SomaScan could be used prospectively to monitor DMD patients. Further method development is needed to validate ART3, as the method we tested was not sufficiently sensitive to reliably detect reduced ART3 levels in serum samples of DMD patients.

Our study has several strengths. We used a well-characterized clinical trial cohort with steroid-naïve patients at baseline and a three year follow-up. Unlike previous studies that focused on a single outcome, we defined biomarkers across multiple functional measures, providing a comprehensive view of patient performance. We also used a reliable high-throughput platform (SomaScan) with low median %CV for screening and identified key biomarkers that were previously reported in two independent cohorts^6^, reinforcing the validity of our findings. However, some limitations should be acknowledged. Serum samples were only available for a subset of patients and time points, reducing the capacity to detect treatment-related effects. Importantly, such selection was not random, therefore the subset of patients with matched serum samples did not replicate the original treatment findings of the FOR-DMD trial.

In conclusion, our data suggests that including a panel of proteins is a viable strategy to monitor DMD patients progression and track treatment response.

## Acknowledgements

The authors thank all patients with Duchenne Muscular Dystrophy and their families for participating in the research study, as well as the study coordinators of FOR-DMD. The collection of the data from the LUMC was funded by Parent Project Muscular Dystrophy through the Protein Mapping Project (Project 22.010). This work was supported by the National Institutes of Health under award number # R61NS119639 (Hathout, Dang, Spitali, Tsonaka, Al-Khalili Szigyarto). This manuscript is the result of funding in whole or in part by the National Institutes of Health (NIH). It is subject to the NIH Public Access Policy. Through acceptance of this federal funding, NIH has been given a right to make this manuscript publicly available in PubMed Central upon the Official Date of Publication, as defined by NIH.

## Supplementary figure

**Figure S1.**
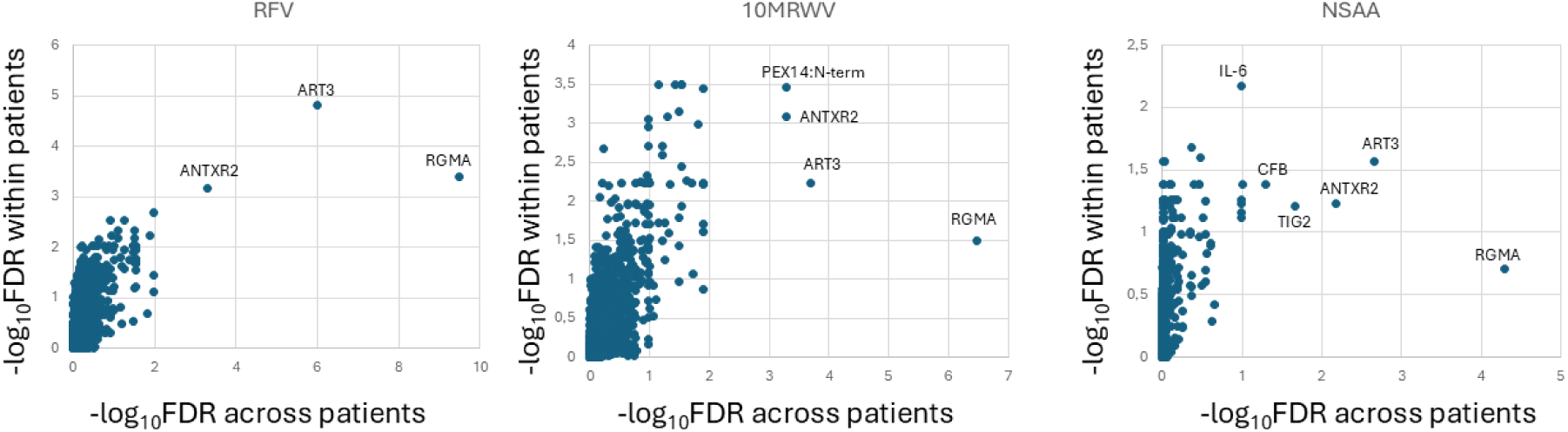
Comparison of the adjusted p-values in the analyses across and within patients.

## Supplementary tables legends

**Table S1.** Protein effect estimates from the across-patient univariate analysis, stratified by physical test and treatment group. Proteins highlighted in red are those associated with the selected physical test and also significantly associated with at least one of the other two tests. SeqId: Somascan aptamer id; Estimates_TREATDAILY_PRED: effect of the protein on the specific outcome for patients treated with daily prednisone; Estimates_TREATDAILY_DFL: effect of the protein on the specific outcome for patients treated with daily deflazacort; Estimates_TREATINT_PRED; effect of the protein on the specific outcome for patients treated with intermittent prednisone; Overall p-value: p-value for the association between functional scores and proteins not corrected for multiple testing; Adjusted p-value: fdr corrected p-value for the association between functional scores and proteins. NSAA: North Star Ambulatory Assessment; RFV: Rise from the Floor Velocity, 10MRWV: 10-Meter Run/Walk Velocity.

**Table S2.** Protein effect estimates from the within-patient univariate analysis, stratified by physical test and treatment group. Proteins highlighted in red are those associated with the selected physical test and also significantly associated with at least one of the other two tests. SeqId: Somascan aptamer id; Estimates_TREATDAILY_PRED: effect of the protein on the specific outcome for patients treated with daily prednisone; Estimates_TREATDAILY_DFL: effect of the protein on the specific outcome for patients treated with daily deflazacort; Estimates_TREATINT_PRED; effect of the protein on the specific outcome for patients treated with intermittent prednisone; Overall p-value: p-value for the association between functional scores and proteins not corrected for multiple testing; Adjusted p-value: fdr corrected p-value for the association between functional scores and protein. NSAA: North Star Ambulatory Assessment; RFV: Rise from the Floor Velocity, 10MRWV: 10-Meter Run/Walk Velocity.

**Table S3.** Percentage of times each of the proteins selected in the univariate analysis (12 in the across-patient analysis and 61 in the within-patient analysis) was chosen by the LASSO model across B = 1000 bootstrap repetitions. SeqId: Somascan aptamer id; Percentage NSAA: percentages from the model with NSAA as the outcome; Percentage 10MRWV: percentages from the model with 10MRWV as the outcome; Percentage RFV: percentages from the model with RFV as the outcome. NSAA: North Star Ambulatory Assessment; RFV: Rise from the Floor Velocity, 10MRWV: 10-Meter Run/Walk Velocity.

